# Retina-derived Quantitative Biomarkers of Brain Health

**DOI:** 10.64898/2026.07.05.26357344

**Authors:** Taizhangtian Ma, Tao Yan, Jing Sun, Ning Wu, Mingze Xu, Ruiheng Zhang, Na Zeng, Qi Sun, Ying Hui, Yuntao Wu, Zhenchang Wang, Tien Yin Wong, Han Lv, Hui Qiao

**Affiliations:** Department of Automation, Tsinghua University, Beijing, 100084, China; Department of Radiology, Beijing Friendship Hospital, Capital Medical University, Beijing, 100050, China; Institute for Brain and Cognitive Sciences, Tsinghua University, Beijing, 100084, China; Beijing National Research Center for Information Science and Technology, Tsinghua University, Beijing, 100084, China; Beijing Visual Science and Translational Eye Research Institute (BERI), Beijing Tsinghua Changgung Hospital, Tsinghua Medicine, Tsinghua University, Beijing, 100084, China; Department of Medical Imaging Technology, School of Medical Technology, Capital Medical University, Beijing, 101300, China; Center for MRI Research, Academy for Advanced Interdisciplinary Studies, Peking University, Beijing, 100871, China; Beijing Tongren Eye Center, Beijing Tongren Hospital, Capital Medical University, Beijing, 100730, China; Department of Infection Control, Peking University First Hospital, Beijing, 100034, China; Precision and Intelligence Medical Imaging Lab, Beijing Friendship Hospital, Capital Medical University, Beijing, 100050, China; Department of Radiology, Kailuan General Hospital, Tangshan, 063000, Hebei Province, China; Department of Cardiology, Kailuan General Hospital, Tangshan, 063000, Hebei Province, China; Department of Health Data Science, School of Medical Technology, Capital Medical University, Beijing, 101300, China; School of Clinical Medicine, Tsinghua Medicine, Tsinghua University, Beijing, 100084, China; School of Biomedical Engineering, Tsinghua Medicine, Tsinghua University, Beijing, 100084, China

## Abstract

Accurate and scalable assessment of quantitative neuroimaging biomarkers, such as white matter hyperintensities (WMH) and hippocampal (HIP) volumes, is essential for understanding and monitoring brain health, preventing neurological diseases and improving healthspan. However, population-level evaluation of these neuroimaging biomarkers relies on inaccessible, costly and time-consuming magnetic resonance imaging (MRI). Here we propose RetiBrain, a cross-modal deep learning framework that predicts these neuroimaging biomarkers from retinal color fundus photography (CFP) images. By distilling latent structural representations from MRI-based models into a CFP-based model, RetiBrain establishes biologically grounded eye-to-brain mapping. In a CFP–MRI paired cohort, RetiBrain accurately estimates six WMH- and HIP-related biomarkers and outperforms the state-of-the-art retinal foundation model RETFound, improving the mean Pearson correlation coefficient by 0.309 (from 0.240 to 0.549) and achieving a coefficient of 0.640 for periventricular WMH prediction. By integrating structural, topological and geometric feature analyses from CFP images, RetiBrain identifies interpretable retinal representations associated with neurodegeneration and cerebrovascular injury, hallmarks of major neurological diseases such as dementia and stroke. In a longitudinal cohort comprising 2,082 participants (4,164 CFP images with up to 15 years of follow-up), RetiBrain-predicted neuroimaging biomarkers robustly estimated neurological disease risk, as illustrated by dementia prediction (AUROC of 0.824, hazard ratio 2.500 per standard deviation increase, 95% CI: 2.201–2.840). RetiBrain provides a robust, scalable, cost-effective and convenient approach for the assessment of neuroimaging biomarkers, and has potential for long-term brain health monitoring in large-scale general population settings.

## Introduction

Brain health represents a dynamic continuum of physiological states, spanning from normal healthy aging and preclinical structural alterations to overt subclinical disease, and ultimately clinically manifest major neurological diseases such as Alzheimer’s disease (AD) and stroke^1^. Thus, lifelong monitoring and maintaining brain health is more than preventing clinically manifest neurological disease^2^. Accurate, convenient and scalable assessment of individual brain health across lifespan is essential for understanding and preventing major neurological diseases, as well as for improving lifelong healthspan^3^.

Currently, precise assessment of brain health status depends on the early detection and evaluation of key preclinical neuroimaging biomarkers that reflect underlying neuropathological changes. For instance, white matter hyperintensities (WMH) are widely recognized as critical indicators of cerebral small vessel disease and vascular cognitive impairment^4–6^, and clinical stroke^7,8^, while hippocampal (HIP) atrophy is a robust hallmark of neurodegenerative progression^9,10^. Currently, magnetic resonance imaging (MRI) represents the gold standard for the detection and quantitative assessment of these neuroimaging biomarkers^11^. However, MRI is expensive, difficult to access in primary care and resource-constrained settings, and the procedure is time-consuming to perform (requiring an average of 35 minutes per brain examination^12^). Thus, routine assessment of WMH and HIP changes remains impractical. The development of a more cost-effective, scalable and accessible alternative for evaluating these neuroimaging biomarkers outside of the MRI suite would facilitate widespread community-based brain health monitoring^13^.

The retina provides a unique opportunity for non-invasive assessment of brain health and neurological diseases. As an extension of the central nervous system^14,15^ with shared embryological origins and physiological connections with the brain, the retina contains structural and microvascular features associated with brain alterations. Previous studies have reported strong associations between retinal changes detected by color fundus photography (CFP) images and neurological diseases, such as stroke^16–24^, cognitive decline^19^ and dementia^25–27^ as well as genetic associations between retinal traits and brain MRI phenotypes^28^, supporting the concept that retinal imaging may serve as an accessible window into brain health and neuropathological processes.

However, the existing deep learning (DL) approaches rely on direct image-to-disease prediction paradigms. Functioning as “black-box” systems, these end-to-end architectures offer limited biological interpretability and fail to elucidate the underlying mechanisms linking retinal features to brain health^29–31^, since disease labels (e.g., AD, stroke) represent final, heterogeneous clinical endpoints rather than quantitative measures of underlying disease process and pathology^32^.

To address these challenges, we introduce RetiBrain, a cross-modal DL framework to estimate neuroimaging biomarkers, principally WMH and HIP, directly from CFP images. Instead of directly predicting neurological outcomes, RetiBrain transfers latent structural representations from MRI into retinal networks through cross-modal distillation (CMD), enabling precise estimation of neuroimaging biomarkers from CFP images. Furthermore, the model integrates structural, topological, and geometric retinal features to enhance predictive performance and establish biologically interpretable links between retinal characteristics and neuroimaging biomarkers, thereby mitigating the “black-box” limitation. To validate this approach, we first evaluated RetiBrain in the Kailuan study and the UK Biobank (UKB) to assess its ability to predict MRI-derived neuroimaging biomarkers from CFP images. The model demonstrated robust and consistent performance across six WMH- and hippocampal-related biomarkers compared with state-of-the-art retinal foundation models. To further examine its clinical relevance, we evaluated whether these non-invasive, CFP-derived biomarkers could support long-term neurological risk stratification in the UKB cohort without requiring

MRI or outcome labels at inference. Across up to 15 years of follow-up, RetiBrain enabled effective identification of individuals at elevated risk of stroke, AD, and dementia, outperforming conventional demographic-based models. These results demonstrate that retinal imaging, augmented by cross-modal representation learning, can serve as a simpler, cost-effective, scalable surrogate for quantitative brain health assessment and enable population-level neurological risk evaluation.

## Results

### Overview of the RetiBrain Framework

The RetiBrain system was trained to reconstruct neuroimaging biomarkers and predict future neurological disease risk using widely available CFP images(Fig. 1a). To bridge the gap between retinal images and brain structures, RetiBrain was built upon a CMD framework that employs an MRI-based teacher network to extract high-dimensional structural features from ground-truth brain MRI (Fig. 1b). These brain embeddings serve as latent supervisory signals, guiding the retinal encoder to align its ocular representations with corresponding neuroimaging features, thereby capturing neuroimaging features within the retinal feature space. The latent structural representations from MRI are transferred into retinal networks with a 3-stage training strategy.

**Fig. 1.**
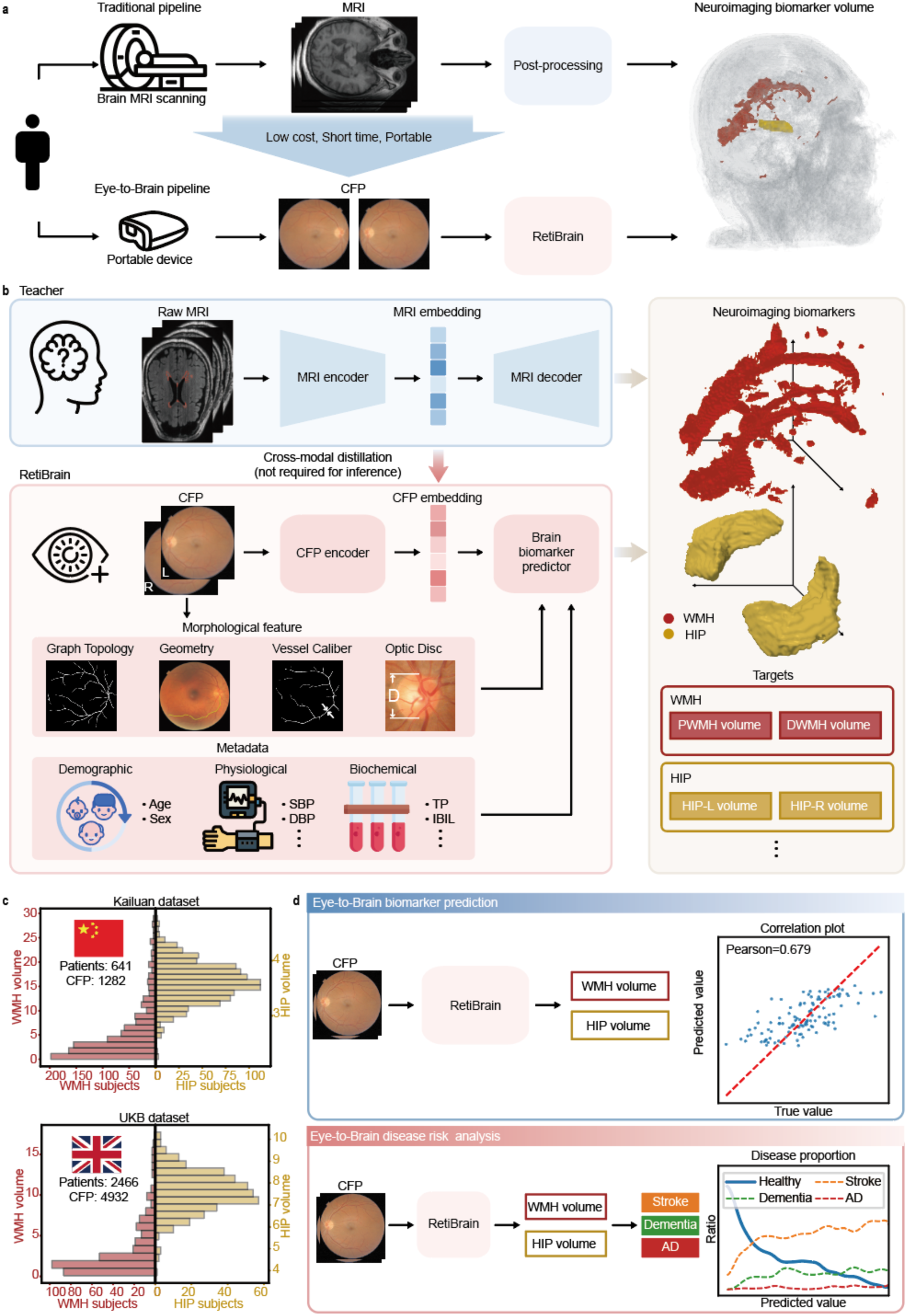
Study design of the present work. a, Comparison of workflows: traditional MRI-based pipeline vs. the proposed Eye-to-Brain pipeline. **b,** A schematic overview of the RetiBrain system architecture. The system integrates retinal images, retinal image-derived morphological features, and individual metadata to predict neuroimaging biomarkers, including white matter hyperintensities (WMH) and hippocampal (HIP) volumes. **c,** Datasets used for model development and validation. A total of 3,107 participants recruited from the Kailuan and UKB datasets were included in this study. The multi-center datasets comprise 6,214 color fundus photograph (CFP) images and 1,025 magnetic resonance image (MRI) scans. **d,** Applications and downstream tasks of the RetiBrain system. Schematic showing the two primary evaluation tracks: Eye-to-Brain biomarker prediction and Eye-to-Brain disease risk analysis.

To optimize continuous biomarker estimation, we augmented these ocular representations with quantitative retinal morphological features. Specifically, the morphological features encompass four distinct domains: (1) graph topology (18 features), characterizing vascular network complexity (e.g., vessel density) to identify localized rarefaction; (2) vascular geometry (27 features), assessing path characteristics such as tortuosity to quantify small-vessel disease severity; (3) vessel caliber (21 features), measuring arterial and venous diameters as indicators of systemic hemodynamics and small-vessel disease burden; and (4) optic disc structure (6 features), evaluating optic nerve head morphometry as a proxy for broader neurodegeneration. Integrating these biologically interpretable features complements the latent representations derived from CFP images by the deep neural network. By synergizing implicit deep-learned features with explicit morphological metrics, RetiBrain more effectively captures retinal pathology.

To further improve model performance, we incorporated demographic and physiological metadata. Demographic variables included age and sex, which represent the fundamental individual-level determinants of brain and retinal health. Physiological variables comprised non-invasive clinical measurements, such as systolic and diastolic blood pressure and body mass index, capturing vascular and metabolic status without requiring blood sampling. We also evaluated the contribution of biochemical variables including blood-derived indicators, such as total protein, albumin and liver function enzymes, providing additional information on systemic metabolic and inflammatory states. By integrating deep-learned ocular representations, explicit retinal morphological features and metadata, RetiBrain is able to better model the eye-brain connection, thereby improving the ability to predict neuroimaging biomarkers.

To establish the clinical validity and generalizability of RetiBrain, we evaluated the framework across the community population-based Kailuan study and the UKB dataset, comprising a total of 6,214 CFP images and 1,025 structural brain MRI (Fig. 1c). Leveraging the multi-center datasets, we implemented a two-stage translational workflow (Fig. 1d). In the first stage, paired CFP–MRI data were utilized to validate the accuracy of RetiBrain in predicting WMH and HIP volumes against MRI-quantified ground truth. In the second stage, we evaluated whether the RetiBrain-predicted biomarkers could stratify the long-term risk of stroke, AD, and dementia in a large UKB subset without MRI data.

### Quantitative Estimation of Neuroimaging Biomarkers in the Kailuan Study

In the Kailuan study, RetiBrain delivered robust quantitative estimation of WMH volumes (total, periventricular [PWMH], and deep [DWMH]) and HIP volumes (total, left [HIP-L], and right [HIP-R]) under a five-fold cross-validation framework (Fig. 2a, b).

**Fig. 2.**
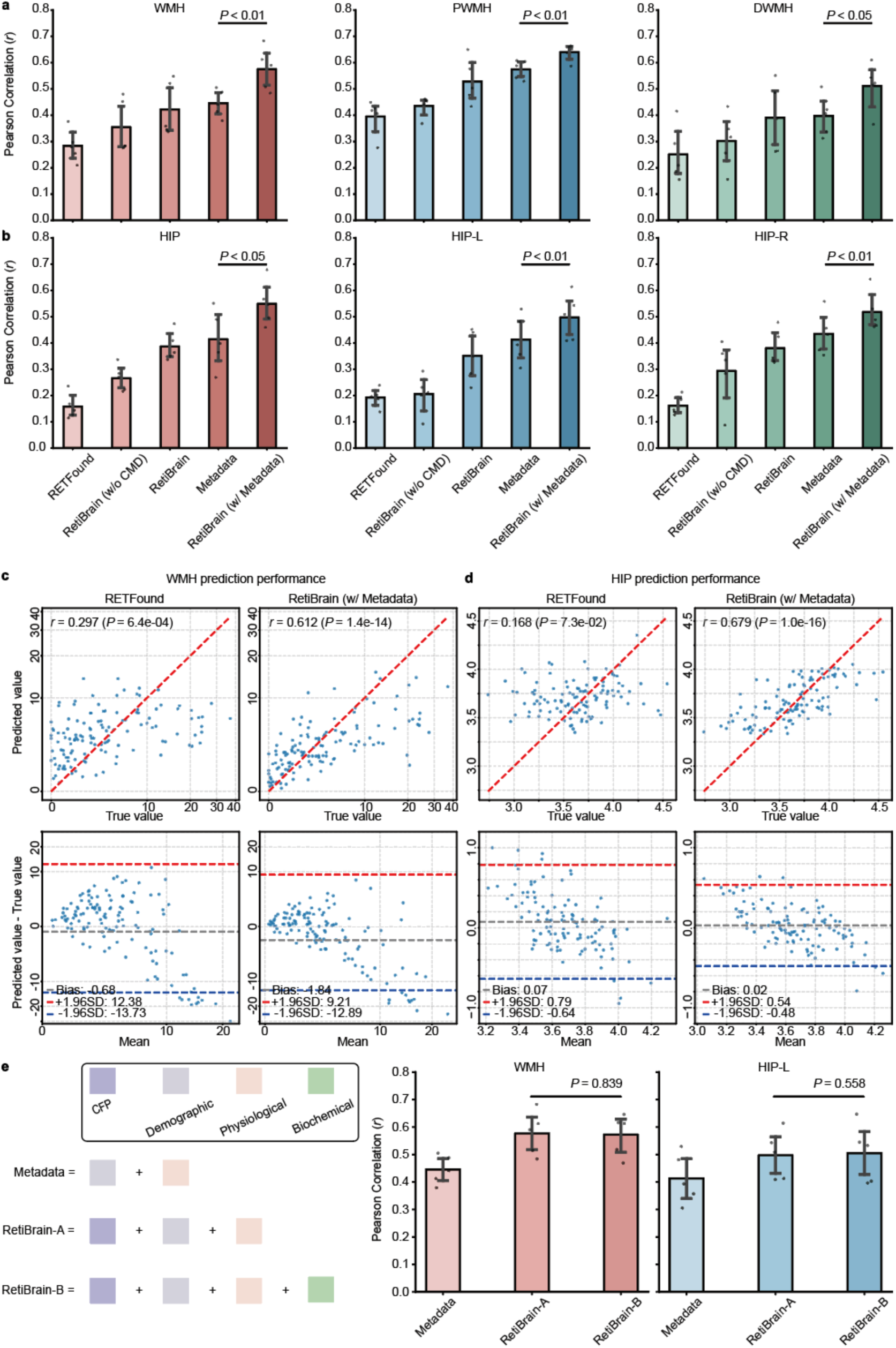
Quantitative Estimation of neuroimaging biomarkers in the Kailuan dataset. a, b, Comparison of Pearson correlation for neuroimaging biomarkers prediction. Bar plots illustrate the performance of 5 models, including RETFound (only color fundus photograph, CFP), RetiBrain (w/o Cross-Modal Distillation, CMD), RetiBrain, Metadata and RetiBrain (w/ Metadata), across the prediction task of WMH and HIP. Morphological features were integrated into all RetiBrain-based variants. Results are presented as mean ± one standard deviation (s.d.) from five-fold cross-validation. Horizontal lines indicate statistical significance between Metadata and RetiBrain (w/ Metadata) (*P* = 0.0022 for WMH, *P* = 0.0014 for PWMH, *P* = 0.415 for DWMH, *P* = 0.0362 for HIP, *P* = 0.0019 for HIP-L, and *P* = 0.0043 for HIP-R; two-sided Student’s t-test). **c, d,** Prediction performance and Bland-Altman analysis for WMH and HIP. The top row shows the correlation between predicted and true values for RETFound and RetiBrain (w/ Metadata). The bottom row displays Bland-Altman plots evaluating the bias and 95% limits of agreement between predicted and real volumes. **e,** Ablation study of input modalities. Left: schematic of input combinations, including Metadata, RetiBrain-A, RetiBrain-B). Right: performance comparison across these configurations for WMH and HIP-L. No statistically significant differences were observed between RetiBrain-A and RetiBrain-B for either WMH (*P* = 0.839) or HIP-L (*P* = 0.558). Statistical significance was assessed using two-sided paired t-test s on paired results from five-fold cross-validation.

Crucially, the full configuration, RetiBrain (w/ Metadata), which integrates CFP images, retinal morphological features, and metadata, including demographic and physiological variables, achieved the best predictive performance, establishing RetiBrain as a robust and accurate framework for continuous neuroimaging biomarkers estimation. Compared with the state-of-the-art retinal foundation model RETFound, RetiBrain (w/ metadata) increased the mean Pearson correlation coefficient (*r*) across all six biomarkers from 0.240 to 0.549, corresponding to an absolute improvement of 0.309, and achieved *r* of 0.640 for PWMH prediction. Analysis revealed that even without the CMD component, the RetiBrain (w/o CMD), which uses CFP images and morphological features, outperformed the RETFound baseline, driving the overall mean *r* of six biomarkers from 0.240 to 0.310. This intermediate improvement indicates that capturing retinal morphological features yields a positive contribution to neuroimaging biomarkers regression. Notably, compared with RetiBrain (w/o CMD), the full CMD framework achieved further statistically significant improvements across most neuroimaging biomarkers ( *P* < 0.05 ), increasing the mean *r* across six biomarkers from 0.310 to 0.410. Relative to RETFound, this corresponds to an absolute improvement of 0.170 (all *P* < 0.05 ). These findings demonstrate that CMD enhances ocular representations for more accurate prediction of neuroimaging biomarkers. We further compared RetiBrain (w/ Metadata) against the Metadata model which makes inference based on demographic and physiological variables. RetiBrain (w/ Metadata) significantly outperformed the Metadata model, elevating the overall mean *r* from 0.446 to 0.549. This result highlights the value of CFP beyond metadata.

Additionally, RetiBrain (w/ Metadata) demonstrated reduced root-mean-square error (RMSE) compared to both the Metadata model and RETFound. These findings confirm that retina provides incremental predictive value for neuroimaging biomarkers, independent of systemic risk factors.

To further visualize and evaluate this predictive performance, we examined the continuous alignment between the inferred values and ground-truth measurements. Compared with RETFound, which achieved modest correlations of *r* = 0.297 for WMH and *r* = 0.168 for HIP, RetiBrain (w/ Metadata) demonstrated substantially improved performance, reaching *r* = 0.612 for WMH and *r* = 0.679 for HIP (Fig. 2c, d). Bland-Altman analysis^33^ confirmed reduced variance in prediction errors (predicted values minus ground-truth values) and narrower limits of agreement.

Finally, an ablation study evaluating the incremental contribution of biochemical profiles revealed a plateau effect: once demographic and physiological variables were incorporated, adding invasive biochemical profiles provided no significant improvement in predictive power (*P* = 0.839 for WMH; *P* = 0.558 for HIP-L; Fig. 2e). Thus, without the need for blood sampling, this approach is highly feasible for large-scale, population-level generalization.

### Interpretability of Brain Biomarker Prediction

Attention heatmaps revealed a shift in the spatial distribution of retinal features associated with increasing brain pathology severity (Fig. 3a). In lower brain pathology cases, model attention was concentrated around the optic disc and its surrounding regions. In contrast, as brain pathology severity increased, attention became more spatially distributed across the fundus, with stronger involvement of the retinal vascular arcades and broader retinal regions. This transition suggests a shift from localized optic disc–centric features toward global retinal structural patterns in higher brain pathology cases, consistent with widespread retinal microvascular remodeling reported in relation to cerebral small vessel disease^21,23,24^.

**Fig. 3.**
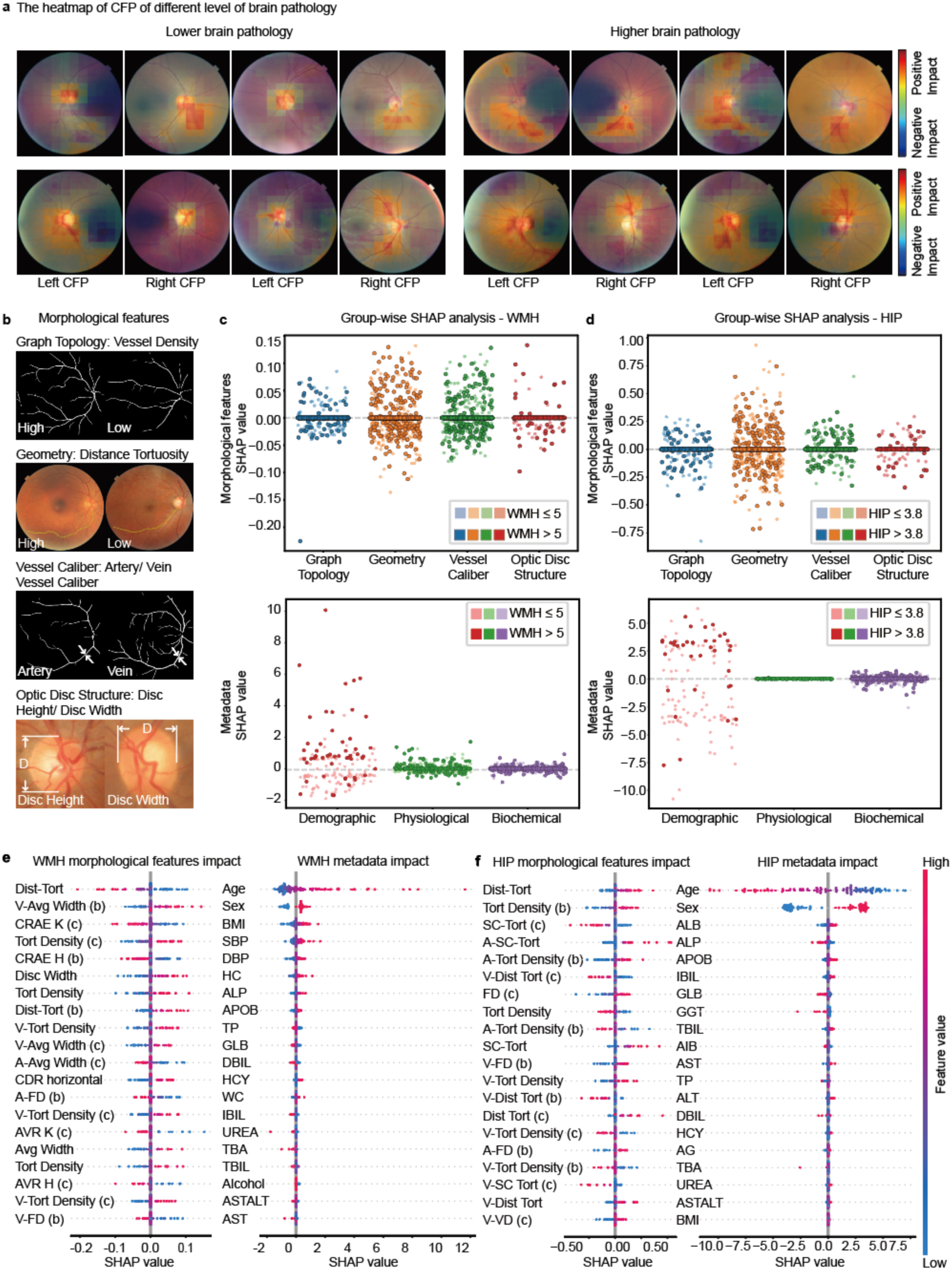
Interpretability of Retinal Features and Metadata for Brain Biomarker Prediction in the Kailuan study. a, Heatmaps for individuals with lower and higher brain pathology are shown. Red regions indicate a positive impact on the predicted biomarker volume, while blue regions indicate a negative impact. **b,** Examples of retinal morphological features. Representative cases illustrate the four feature categories: graph topology, geometry, vessel caliber, and optic disc structure. **c, d,** Group-level feature importance based on SHAP values. Distribution of SHAP values for WMH (c) and HIP (d) are grouped by morphological features and metadata (categorized into Demographic, Physiological, and Biochemical). **e, f,** SHAP summary plots for individual features. Detailed impact of specific morphological features and metadata on the prediction of WMH (e) and HIP (f). Each point represents an individual sample; the color gradient reflects the feature value (red for high, blue for low), and the x-axis position indicates the SHAP value. Abbreviations in **e** and **f**: Dist-Tort, distance tortuosity; Avg Width, average width; CRVE/CRAE, central retinal vein/artery equivalent; V/A, vein/artery; Tort Density, tortuosity density; FD, fractal dimension; VD, vessel density; AVR, artery-vein ratio; SC-Tort, square curvature tortuosity; Disc Width, optic disc width; CDR horizontal, horizontal cup-to-disc ratio; (b), Zone B; (c), Zone C; K, Knapp’s formula; H, Hubbard’s formula. BMI, body mass index; SBP/DBP, systolic/diastolic blood pressure; WC/HC, waist/hip circumference; Age, chronological age; Sex, biological sex; Alcohol, alcohol consumption status; APOB, apolipoprotein B; ALP, alkaline phosphatase; ALB, albumin; GLB, globulin; AIB, albumin in buffer; AG, albumin/globulin ratio; IBIL/DBIL/TBIL, indirect/direct/total bilirubin; HCY, homocysteine; ALT/AST, alanine/aspartate aminotransferase; ASTALT, AST/ALT ratio; GGT, gamma-glutamyl transferase; TP, total protein; TBA, total bile acids; UREA, blood urea.

To bridge qualitative visual patterns with quantifiable structural characteristics, we analyzed 72 high-dimensional retinal morphological features spanning four domains: graph topology, vascular geometry, vessel caliber, and optic disc structure (Fig. 3b). SHAP-based global feature attribution identified vascular geometry as the most influential domain for both WMH and HIP regression tasks (Fig. 3c, d). At the individual-feature level (Fig. 3e, f), distance tortuosity (Dist-Tort) emerged as the predominant predictor for both WMH and HIP, exhibiting the highest feature attribution across both targets. Notably, the prominent contribution of vascular tortuosity aligns with existing longitudinal cohort evidence linking retinal vascular tortuosity to subsequent dementia risk^25,27^.

Regarding metadata, demographic factors, particularly chronological age, exerted the largest global effects across both WMH and HIP targets (Fig. 3c, d). Age was positively associated with high-burden WMH but negatively associated with hippocampal volume. For individual metadata features in WMH prediction, body mass index (BMI), systolic blood pressure (SBP), and diastolic blood pressure (DBP) emerged as the primary drivers^34^ (Fig. 3e). In contrast, HIP volume prediction predominantly relied on albumin (ALB) and alkaline phosphatase (ALP) levels (Fig. 3f). These findings suggest that WMH may be associated with chronic systemic vascular stress, while hippocampal atrophy may be more closely correlated with metabolic and biochemical dysregulation.

### Prediction of Neuroimaging Biomarkers and Neurological Diseases in the UK Biobank with Paired CFP-MRI Data

To validate the generalizability of RetiBrain, we further evaluated RetiBrain’s cross-modal biomarker estimation capabilities in a subset of UKB with paired CFP-MRI data. RetiBrain accurately predicted neuroimaging biomarkers from retinal images (Fig. 4a), achieving Pearson correlation coefficients with ground-truth MRI measurements for PWMH (*r* = 0.690) and HIP (*r* = 0.476). Subgroup analysis indicated significant differences in the predicted volumes of PWMH and HIP between participants who subsequently developed stroke and dementia and those who remained disease-free (Fig. 4b).

**Fig. 4.**
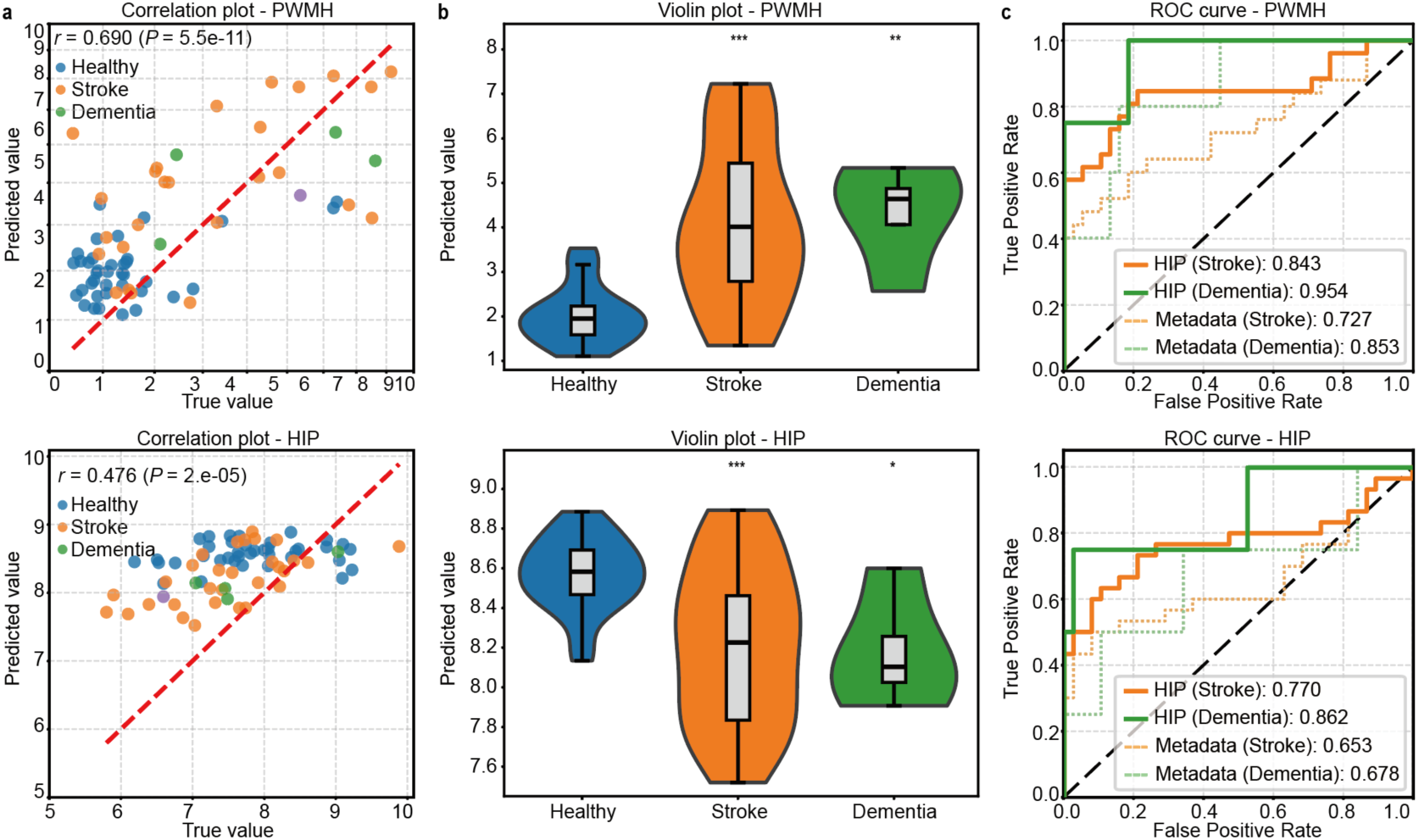
Prediction of neuroimaging biomarkers and neurological diseases in the UK Biobank with paired CFP-MRI data. a, Correlation analysis of predicted versus true biomarkers. Scatter plots illustrate the prediction performance of RetiBrain for PWMH and HIP volumes within the UKB subset with available MRI ground truth. **b,** Distribution of predicted biomarkers across clinical groups. Violin plots show the variation in predicted PWMH and HIP volumes among healthy, stroke, and dementia participants. Significant differences between the healthy control group and disease groups were determined using the two-sided Mann-Whitney U test. Statistical significance is indicated as: * *P* < 0.05 ; ** *P* < 0.01 ; *** *P* < 0.001 . **c,** Comparison of classification performance between RetiBrain and metadata for stroke and dementia. 95% confidence intervals were estimated via bootstrapping with 1000 resamples.

We further assessed whether RetiBrain-predicted biomarkers derived from baseline CFP images could discriminate individuals who subsequently developed disease using receiver operating characteristic (ROC) analyses (Fig. 4c). Predicted PWMH identified future dementia with an area under the receiver operating characteristic curve (AUC) of 0.954 (95% confidence interval, CI: 0.829–1.000), representing an AUC increment of 0.101 over the metadata-only model. Collectively, these results demonstrate that RetiBrain captures critical brain health-related features beyond common metadata, enabling more accurate risk stratification for neurological diseases.

### Neurological Disease Risk Stratification in the UK Biobank with CFP-only Data

To translate the eye-brain associations established in the paired UKB cohort into clinical utility, we evaluated whether RetiBrain-predicted biomarkers could generalize to large-scale populations lacking brain MRI data and retaining prognostic fidelity for future neurological diseases. To reduce potential confounding from differences in age distribution between healthy individuals and those who subsequently developed neurological diseases, we analysed a CFP-only UKB subset with similar age distributions across groups (Fig. 5a). RetiBrain-predicted PWMH and HIP volume exhibited highly significant differences between individuals who remained healthy and those who subsequently developed neurological diseases (*P* < 0.05; Fig. 5b). The results confirm that RetiBrain captures robust neurodegenerative signals from retinal structures, independent of ground-truth brain MRI.

**Fig. 5.**
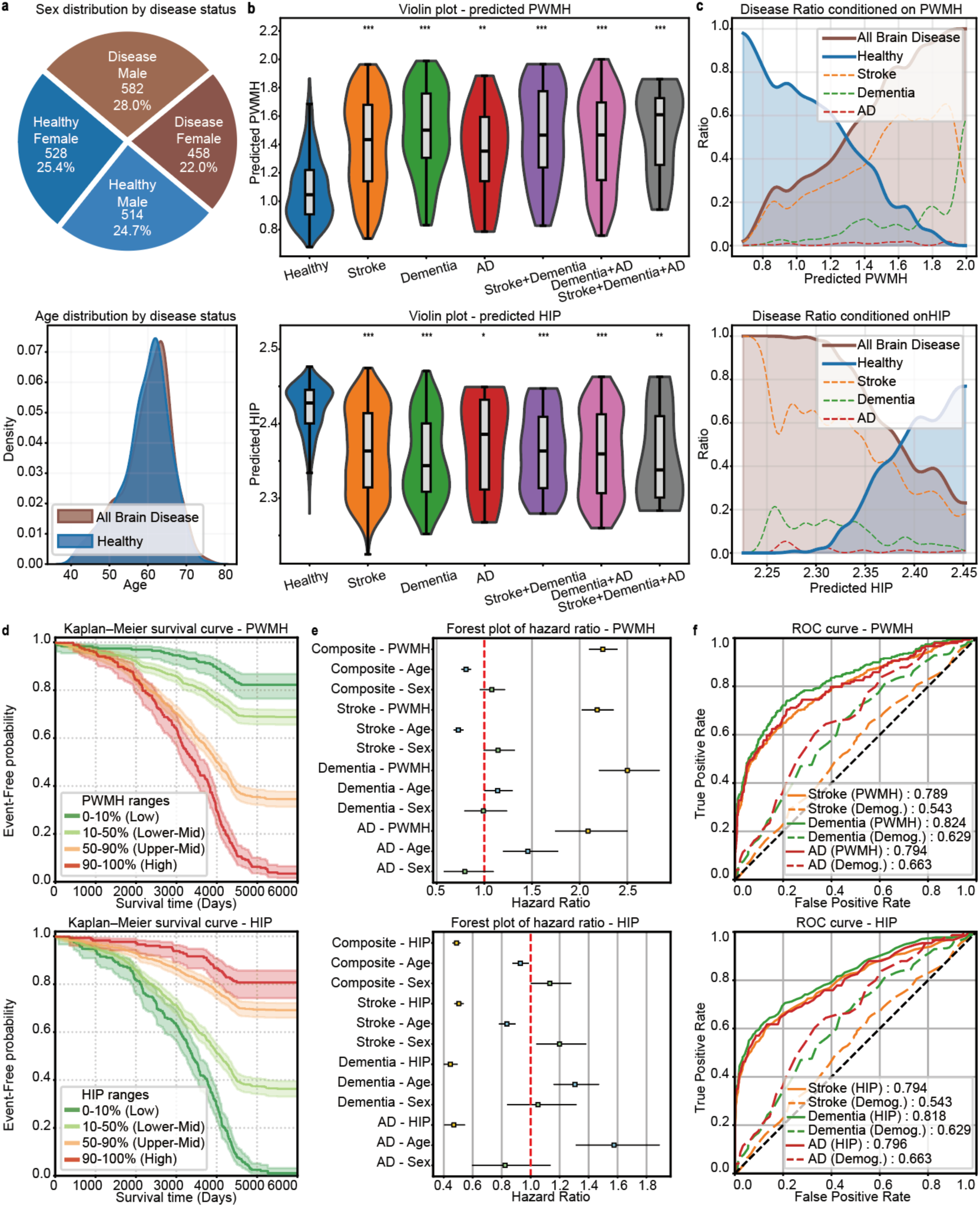
Longitudinal neurological risk stratification using RetiBrain-predicted neuroimaging biomarkers in the UK Biobank with CFP-only data. a, Demographic characteristics of the UKB cohort in CFP-only setting. The distributions of age and sex are shown for healthy individuals and subjects with neurological diseases. **b,** Predicted biomarker distribution across disease phenotypes. Violin plots display the variation in RetiBrain-predicted PWMH and HIP volumes among healthy individuals and those who subsequently developed stroke, dementia, Alzheimer’s disease (AD), or comorbid conditions. Significant differences between the healthy control group and disease groups were determined using the two-sided Mann-Whitney U test. Statistical significance is indicated as: \**P* < 0.05; ** *P* < 0.01; *** *P* < 0.001. **c,** Disease ratio conditioned on predicted biomarker levels. The relative proportions of healthy individuals versus those with neurological diseases are plotted across predicted biomarkers. **d,** Kaplan-Meier survival analysis for incident neurological diseases. Cumulative event-free probabilities for the endpoint are stratified by four biomarker ranges (0–10%, 10–50%, 50–90%, and 90–100% percentiles). **e,** Hazard ratios for independent risk factors. Forest plots display the hazard ratio (HR) and 95% confidence intervals for the predicted biomarkers, age, and sex across multiple endpoints, including the composite endpoint (composite), stroke, dementia, and AD. **f,** Predictive performance for future clinical outcomes. ROC curves compare the predictive accuracy of RetiBrain-predicted biomarkers (solid lines) against demographic baselines (Demog.; dashed lines) for future neurological disease risk. AUC was utilized as the metric.

In order to determine whether the RetiBrain-predicted biomarkers enable dose-dependent risk stratification, we quantified disease incidence across the full spectrum of predicted biomarkers (Fig. 5c). Higher predicted PWMH burden was associated with a progressive increase in disease incidence, while lower predicted HIP volumes were correlated with higher disease incidence.

We further conducted longitudinal survival analyses over a 15-year follow-up period (Fig. 5d). Stratification into percentile-based cohorts revealed divergent disease trajectories over time. The highest-percentile PWMH and lowest-percentile HIP groups showed steep declines in event-free probability, while opposite percentiles remained stable. By year 15, nearly all individuals in the highest-risk percentiles progressed to clinical disease, compared with approximately 80% event-free in the healthiest tiers.

Multivariate Cox regression further established the independent prognostic value of RetiBrain-predicted biomarkers beyond metadata (Fig. 5e). We analyzed the impact of age, sex, and biomarkers. Each standard deviation (s.d.) increase in predicted PWMH was associated with a 2.500-fold higher dementia risk (hazard ratio, HR = 2.500; 95% CI: 2.201–2.840), an effect substantially exceeding that of age (HR = 1.139; 95% CI: 1.001–1.297) and sex (HR = 0.990; 95% CI: 0.790–1.241). Conversely, each s.d. increase in predicted HIP was strongly protective for developing dementia (HR = 0.445; 95% CI: 0.400–0.496).

Biomarker-only models enabled prediction of multiple neurological outcomes, including dementia, AD, and stroke (Fig. 5f). For dementia, the models achieved high accuracy, yielding an AUC of 0.824 (95% CI: 0.795–0.851) for predicted PWMH and 0.818 (95% CI: 0.786–0.845) for predicted HIP. Notably, these biomarker-derived models substantially outperformed demographic-only baselines, with absolute AUC gains of 0.195 and 0.189, respectively. Across other neurological endpoints, RetiBrain showed strong predictive performance for both AD and stroke, with PWMH- and HIP-derived models achieving AUCs of 0.794 (95% CI: 0.773–0.816) and 0.796 (95% CI: 0.750–0.838) for AD, and 0.789 (95% CI: 0.768–0.810) and 0.794 (95% CI: 0.773–0.816) for stroke, respectively.

These findings were further supported by comprehensive analyses across other RetiBrain-predicted biomarkers, including WMH, DWMH, HIP and HIP-R, with consistent results observed in group comparisons, risk stratification, longitudinal survival analyses, multivariate Cox regression and disease prediction. Collectively, these findings establish RetiBrain as a scalable, non-invasive tool for the detection of preclinical brain structural changes, facilitating the quantitative tracking of long-term brain health trajectories.

## Discussion

There is demand for assessment of brain health with simple, convenient, and cost-effective approaches. Such approaches will allow understanding and lifelong monitoring of brain health, allowing optimal risk stratification and prevention of major clinical neurological diseases such as AD and other dementia, stroke and cerebrovascular diseases, ultimately reducing neurological disease-related morbidity, disability and mortality across populations. We developed RetiBrain, a DL system that leverages CFP images to quantitatively assess key subclinical neuroimaging biomarkers of WMH burden and HIP volume. Using two multicenter, cross-ethnic cohorts comprising participants from China and the UK, we trained and validated RetiBrain. The primary findings demonstrate that RetiBrain achieved strong quantitative agreement with MRI-derived ground truth. Compared with the state-of-the-art retinal foundation model RETFound, RetiBrain substantially improved predictive performance, increasing the mean Pearson correlation coefficient across six biomarkers by an absolute margin of 0.309 (from 0.240 to 0.549), and achieving a coefficient of 0.640 for periventricular WMH. The model achieves robust predictions using only CFP alongside basic demographic and physiological data, bypassing the need for invasive biochemical tests. Interpretability analyses confirmed that RetiBrain relies on biologically grounded features, identifying retinal vascular tortuosity as a pivotal biomarker. Moreover, predicted PWMH burden showed a strong association with dementia risk, yielding a HR of 2.500 per standard deviation increase. Accurate estimation of PWMH and HIP volumes further improved risk prediction for 15-year incident dementia, AD and stroke, with AUC values of 0.824, 0.794, and 0.789, respectively. Notably, RetiBrain demonstrated markedly enhanced risk discrimination compared with conventional clinical risk models based on age and other metadata.

While previous studies have explored the application of CFP images using DL techniques for the assessment of diverse neurological diseases, most existing works directly utilize the entire retinal images to predict categorical disease outcomes (presence or absence of dementia or stroke), which fails to fully exploit the intermediate pathway and subclinical neuropathological alterations. Our present study proposes a novel advancement in research design. It is worth emphasizing that our work aims to not only predict quantitative neuroimaging biomarkers with the proposed RetiBrain system, but also to assess risk of clinical neurological diseases based on RetiBrain-predicted biomarkers. Our strategy further suggests the ability of the retina to be a surrogate marker of the brain, offering novel approaches for neurological diseases prevention and personalized brain health risk management.

From the perspective of model transformation for clinical implementation, adopting these quantifiable and pathophysiological validated neuroimaging features as intermediate biomarkers improves model interpretability and enhances its compatibility with routine clinical practice, thereby substantially boosting clinicians’ trust in AI-assisted diagnosis^35,36^. Prior evidence has validated retinal imaging features as potential biomarkers for neurological diseases^29,37^, while subclinical imaging manifestations on brain MRI, such as WMH lesions and cerebral atrophy, provide direct and objective radiological evidence for disease diagnosis^38–40^. Although cutting-edge DL-based AI

models have achieved outstanding performance in neurological diseases classification, their inherent “black-box” decision mechanisms compromise clinical credibility and impede practical deployment into clinical diagnostic workflows^29–31^. RetiBrain provides traceable biomarker-level explanations, which allows clinicians to cross-verify model predictions against observable neuroimaging findings. This approach effectively transforms the role of AI from AI alone to “human-AI collaborative” model that is important for patient-centered care^35,41^.

Several limitations of this study warrant acknowledgment. First, additional multi-ethnic cohorts with heterogeneous demographics are essential to verify model robustness and generalizability. RetiBrain links retinal phenotypes to neuroimaging biomarkers and neurological diseases, but paired retinal-neuroimaging datasets remain scarce. These datasets are difficult to acquire because neuroimaging is costly and requires specialized infrastructure. To address this challenge, we must collaborate with major clinical institutions to share data and build multi-center validation repositories. Second, further incorporation of multimodal retinal imaging including optical coherence tomography (OCT) may facilitate more comprehensive mapping of eye-brain correlated features. Future work will extend this architecture into a multi-branch network to fuse superficial fundus features with deep metrics from OCT. Finally, the present study utilized baseline brain imaging data. Future iterations will leverage longitudinal cohorts, utilizing sequence-modeling frameworks (e.g., recurrent networks) on serial retinal inputs to dynamically predict the progressive burden of subclinical imaging markers and track clinical disease progression over time.

In conclusion, we developed RetiBrain, a CFP image-based system for the quantitative assessment of subclinical neuroimaging biomarkers, including WMH burden and HIP atrophy. These RetiBrain-predicted biomarkers further enhance risk stratification for future neurological diseases including dementia, AD and stroke. Our work establishes retinal imaging as a scalable, cost-effective and convenient tool for long-term monitoring of brain health and neurological disease prediction and risk stratification at the population level.

## Data availability

The Kailuan dataset used in this study is not publicly available. Access is available to qualified research groups whose proposed use of the data has been reviewed and approved by an independent review committee established for this purpose. Data access requests may be directed to Dr. Zhen-Chang Wang. The UK Biobank data that support the findings of this study are available to approved researchers upon application via the UK Biobank access procedure at https://www.ukbiobank.ac.uk/.

## Code availability

The code can be found at https://github.com/MTZT2002/RetiBrain/

## Data Availability

The Kailuan dataset used in this study is not publicly available due to participant privacy protection and institutional data-use restrictions. Access may be available to qualified research groups whose proposed use of the data has been reviewed and approved by an independent review committee established for this purpose. Data access requests may be directed to Dr. Zhen-Chang Wang. The UK Biobank data that support the findings of this study are available to approved researchers upon application through the UK Biobank access procedure. The UK Biobank data that support the findings of this study are available to approved researchers upon application through the UK Biobank access procedure at https://www.ukbiobank.ac.uk/.

## Acknowledgments

This work was supported by the National Natural Science Foundation of China (grant nos. 62522119, 82502453 and 62322110), the National Key R&D Program of China (grant no. 2023YFB3209700), the China Postdoctoral Science Foundation (grant nos. 2025M782188, BX20240196 and 2025M780784), the R&D Program of Beijing Municipal Education Commission (grant no. KM202410025017), and the Shuimu Tsinghua Scholar Program.

## Author contributions

H.Q. and H.L. conceived and supervised the study. T.Y., T.M., J.S., H.Q., and H.L. designed the methodology. T.M., T.Y., and J.S. implemented the algorithms and analyzed the results. H.L., N.W. and M.X. contributed to the data acquisition, organization, and preprocessing. R.Z., N.Z., Q.S., Y.H., Y.W., and Z.W. provided the clinical inputs and contributed to the evaluation pipeline. T.M., T.Y., J.S., H.L. and T.W. prepared the manuscript with input from all the authors. All authors contributed to the review and final revision of the manuscript.

